# Diffusion-based structural connectivity patterns of multiple sclerosis phenotypes

**DOI:** 10.1101/2023.03.21.23287029

**Authors:** E Martinez-Heras, E Solana, F Vivó, E Lopez-Soley, A Calvi, S Alba-Arbalat, MM Schoonheim, EMM Strijbis, H Vrenken, F Barkhof, MA Rocca, M Filippi, E Pagani, S Groppa, V Fleischer, R Dineen, B Ballenberg, C Lukas, D Pareto, À Rovira, J Sastre-Garriga, S Collorone, F Prados, AT Toosy, O Ciccarelli, A Saiz, Y Blanco, S Llufriu

## Abstract

**Background:** We aimed to describe the severity of the changes in brain diffusion-based connectivity as multiple sclerosis (MS) progresses and the microstructural characteristics of these networks that are associated with distinct MS phenotypes.

**Methods:** Clinical information and brain magnetic resonance images were collected from 221 healthy individuals and 823 people with MS at eight MAGNIMS centers. The patients were divided into four clinical phenotypes: clinically isolated syndrome, relapsing-remitting, secondary-progressive, and primary-progressive. Advanced tractography methods were used to obtain connectivity matrices. Then, differences in whole-brain and nodal graph-derived measures, and in the fractional anisotropy of connections between groups were analyzed. Support vector machine algorithms were used to classify groups.

**Results:** Clinically isolated syndrome and relapsing-remitting patients shared similar network changes relative to controls. However, most global and local network properties differed in secondary progressive patients compared with the other groups, with lower fractional anisotropy in most connections. Primary progressive participants had fewer differences in global and local graph measures compared to clinically isolated syndrome and relapsing-remitting patients, and reductions in fractional anisotropy were only evident for a few connections. The accuracy of support vector machine to discriminate patients from healthy controls based on connection was 81%, and ranged between 64% and 74% in distinguishing among the clinical phenotypes.

**Conclusions:** In conclusion, brain connectivity is disrupted in MS and has differential patterns according to the phenotype. Secondary progressive is associated with more widespread changes in connectivity. Additionally, classification tasks can distinguish between MS types, with subcortical connections being the most important factor.

**What is already known on this topic:**

- MS is a neurodegenerative disease characterized by inflammation and demyelination in the central nervous system, leading to disrupted neural connections and varying clinical phenotypes.
- Diffusion-based MRI techniques and graph theory can be used to study microstructural changes and brain network alterations in MS patients across different phenotypes.

**What this study adds:**

- The study highlights distinct patterns of brain connectivity disruptions associated with different MS phenotypes, particularly revealing more widespread changes in connectivity for secondary-progressive MS.
- It demonstrates the effectiveness of support vector machine algorithms in classifying patients from healthy controls (81% accuracy) and distinguishing among clinical phenotypes (64% to 74% accuracy) based on brain connectivity patterns.
- The study emphasizes the importance of subcortical connections as a key factor in differentiating MS types, providing valuable insights into the underlying neural mechanisms related to MS phenotypes.

**How this study might affect research, practice or policy:**

- This study might affect research, practice, or policy by providing a better understanding of the differential patterns of brain connectivity disruptions across MS phenotypes, which can guide the development of more accurate diagnostic and prognostic tools, leading to improved personalized treatment and management strategies for people with multiple sclerosis.

## INTRODUCTION

Multiple sclerosis (MS) is a chronic inflammatory, demyelinating and neurodegenerative disorder of the central nervous system, which is characterised by varying degrees of physical and cognitive disability ^1^. The evolution of this disease is very heterogeneous: patients with a clinically isolated syndrome (CIS) commonly progressing to a relapsing-remitting (RRMS) disease course and eventually, experiencing a poorly understood sustained deterioration in their disability independent of relapses after several years, referred to as secondary progressive MS (SPMS). In addition, a smaller percentage of patients develop a progressive course from the very beginning, known as primary-progressive MS (PPMS) ^2^.

MS is characterised by the presence of focal and diffuse damage within the white and grey matter of the brain ^3^, which accumulates as the disease progresses and disrupts structural connections in the brain ^4^. Diffusion magnetic resonance imaging (MRI) can offer information regarding the integrity of white matter (WM) connections, representing the topology and hierarchy of structural brain networks ^5^. The disconnection among MS brain networks is characterised by impaired information flow and worse network efficiency, mainly due to poor integration between cortical areas and the enhanced segregation of brain regions ^6^. In particular, long-range anatomical tracts are damaged, and at later stages of the disease the disruption of the connections between network hubs becomes more prominent ^7^. The changes in networks are quite heterogeneous in MS, depending mainly on the areas damaged and the severity of the disruptions ^8,9^. In fact, compensatory reorganization can to some extent preserve the global efficiency of the brain in the early stages of the disease, mainly manifested by functional changes ^10^. By contrast, at more advanced stages of MS structural damage leads to less efficient network wiring and when this loss of efficiency reaches a critical threshold, the network collapses and clinical progression accelerates ^11^.

At present, the severity and characteristics of the modifications to structural connectivity associated with MS phenotypes remain poorly understood. Although previous studies have shown significant differences in global and local network metrics between RRMS and SPMS ^12^, modelling the disconnection caused by lesions did not result in significant changes in connectivity. Hence, it would appear that those previous results of an overall decline in network integrity were caused principally by the effects of lesion load on the underlying tissue structure. Preliminary studies that employed machine learning (ML) algorithms and binary classifications to discriminate clinical phenotypes based on the characteristics of connectivity displayed good accuracy, suggesting that the distinct MS phenotypes are associated with different network modifications ^13,14^, but this awaits confirmation in larger samples.

Here, we set out to comprehensively test the clinical relevance of structural network changes in MS by describing the modifications to brain networks and their components associated with the different MS phenotypes, and to assess the ability of these modifications to classify the changes in connectivity. To this end, we studied a large cohort of people with MS (PwMS) recruited through a multicentre collaboration within the MAGNIMS network.

## METHODS

### Participants

In this retrospective cross-sectional study, we included data from eight centres across Europe that were members of the MAGNIMS network (https://www.magnims.eu/): [1] ImaginEM group at the Hospital Clinic Barcelona (Spain); [2] The Amsterdam MS Center, Amsterdam UMC, location VUmc, Amsterdam (The Netherlands); [3] The Neuroimaging Research Unit, San Raffaele Scientific Institute, Milan (Italy); [4] University Medical Center of the Johannes Gutenberg University Mainz (Germany); [5] Division of Clinical Neuroscience, University of Nottingham, Nottingham (UK); [6] St. Josef Hospital Ruhr University, Bochum (Germany); [7] The Cemcat and Section of Neuroradiology, University Hospital Vall d’Hebron, Barcelona (Spain); and [8] Institute of Neurology, UCL, London (UK). None of the participants had any clinically signs of relapse nor had they received steroid treatment in the 30 days prior to the study visit. After a quality-control check, the clinical and demographic information from the 823 PwMS and 221 healthy controls (HCs) was analyzed, as were the MRIs acquired from these participants between 2011 and 2019.

The ethical review board at the Hospital Clinic in Barcelona approved the study and all the participants gave their written consent for their data to be used. Data transfer agreements were established with the participating centres in order to allow sharing of pseudonymized images and clinical information governed by a central MAGNIMS collaboration framework agreement.

### Magnetic resonance acquisition and processing

MRI scans were collected for each participant site using 3T scanners (different vendors) using common MRI acquisition parameters (MRI details for each centre are provided in Supplementary Table 1: a) 3D-T1 structural sequences with ≤ 1.5 mm isotropic voxel size; and b) whole brain diffusion scans with at least 30 diffusion encoding directions, ≤ 2.5 mm isotropic voxel size and b-value ≥ 900 s/mm^2^. We computed whole brain and local (nodal) graph metrics, as well as the fractional anisotropy (FA) levels from each connection contributing to the network.

### Anatomical and diffusion processing pipelines to build the structural brain networks

Along with the MRI data, all the centres involved in this study provided the corresponding white matter lesion segmentation mask for each subject. These masks were used to obtain lesion-filled T1w images in order to guarantee accurate segmentation ^15^. We estimated global brain and lesion volumes with FSL-SIENAX software (https://fsl.fmrib.ox.ac.uk/fsl/fslwiki/SIENA) and harmonized these values through ComBat, including as a covariate MS phenotypes, to protect against the influence of different center sites ^16^. The lesion-filled T1w images were parcellated into 62 cortical and 14 subcortical grey matter (GM) regions using the anatomical Desikan-Killany atlas (as generated with Mindboggle software from T1w images processed by ANTs cortical thickness and through the FSL-FIRST pipeline), and these regions were considered the nodes of the network ^17,18,^.

To reduce the confounding effects derived from diffusion-weighted images (DWI) acquisition, a well-established DWI preprocessing pipeline was followed using the FSL and MRtrix packages, as described previously ^19^. These preprocessing steps involved skull stripping, Gibbs ringing correction, MP-PCA denoising, slice-wise outlier detection, eddy current and motion correction within DWI volumes (using FSL’s eddy tool), rotation of the b-vectors, geometric distortion corrections depending on each dataset involved (gradient field maps, performing a non-linear registration of the b0 image to an undistorted T2 anatomical image or using a B0 undistorted synthetic image) ^20,21^, and field bias correction. After these corrections, model fitting was performed on the undistorted DWI to estimate the diffusion tensor and then compute the quantitative FA scalar map using FSL’s DTIFIT ^22^. In addition, the undistorted b0 images were used to improve the accuracy of DWI registration to anatomical T1w images.

Quantitative FA connectivity matrices were obtained by performing constrained-spherical deconvolution (CSD) and probabilistic advanced diffusion tractography ^23^. A set of 6 million streamlines were generated into WM mask to capture the entire WM fibre trajectories, which were established by applying anatomically constrained tractography (5-tissue-types) ^24^ and 2850 WM connections assessed, according to all pairs of cortical and deep grey matter (dGM) regions. Subsequently, anatomical exclusion criteria were applied to reduce the number of spurious streamlines from each connection ^25^. We defined pairs of GM regions as structurally connected if the connection was preserved after applying SIFT2 functions ^26^. The mean FA along each reconstructed fibre pathway was computed to establish the whole-brain structural connectome between all pairs of GM regions. Finally, the connections that were present in less than 60% of the HCs group were removed from the network, ending up with 1818 connections, and we then corrected the effects of age and gender using linear regression ^27^. As the FA-weighted adjacency matrices could suffer from inter-site variability related to heterogeneity in the acquisition protocol, we harmonized the data with the ComBat model using an imputation technique for missing values ^28^.

### Network metrics

Global and local network graph measures were estimated using the Brain Connectivity toolbox (https://sites.google.com/site/bctnet/) ^29^. We investigated local network measures like nodal strength (the sum of the edge weights connected to a node), nodal betweenness-centrality (the number of the shortest paths that pass through a node), nodal clustering coefficient (the fraction of the node’s neighbours that are closed as triangles) and nodal local efficiency (the inverse of the shortest path distance between the nodes). In addition, we calculated the average of these local topological features for all the nodes involved in each network and the global efficiency to measure the global network properties.

### Statistical analyses and classification tasks

A two-sample Student’s t-test was carried out to assess differences in the network metrics and FA connectivity matrices between HCs and PwMS, as well as among different MS phenotypes. We applied multiple comparison tests with the Benjamini/Yekutieli method to control for false positives (*p* < 0.05) and only variables with an effect size, measured using Cohen’s D, greater than 0.6 in absolute value were considered for further statistical analysis.

We used a support vector machine (SVM) classification algorithm to discern between the HCs and MS groups, and to classify the disease phenotypes using binary classification tasks once the significant graph-derived measures and the FA-weighted connections had been selected from the group differences. For this task, we analysed the Receiver Operating Characteristic (ROC) dealing with unbalanced data (sub-sampling the groups with a larger number of subjects, CIS/RRMS) to predict the classification accuracy (F-measure) among MS subtypes through 10-fold cross-validation over the entire dataset selected. These models were also investigated using confusion matrices to assess other indicators of the classification’s performance, such as precision, sensitivity and specificity. Finally, the 20 most important inter-regional connections were identified in each machine learning model proposed.

## RESULTS

The clinical and demographic information was collected from the 823 PwMS (74 CIS, 571 RRMS, 121 SPMS and 57 PPMS) and 221 HCs included in the study. As expected, the PwMS with progressive forms of the disease were older and had higher scores on the Expanded Disability Status Scale (EDSS), as well as a longer disease duration (*p* < 0.05). Lesion volume was larger in SPMS than in the other phenotypes, while normalized brain volume was smaller in SPMS and PPMS compared with the CIS and RRMS groups (Table 1).

**Table 1.** Characteristics of the sample.

|  | **HCS**<br>**(n= 221)** | **CIS**<br>**(n= 74)** | **RRMS**<br>**(n= 571)** | **SPMS**<br>**(n= 121)** | **PPMS**<br>**(n= 57)** | **p-value**<br>**HCs vs PwMS** | **p-value**<br>**MS types** |
| --- | --- | --- | --- | --- | --- | --- | --- |
| Age, median in years (range) | 41.2<br>(30.3-49.7) | 34.1<br>(28.9-42.3) | 43<br>(35.9-50.6) | 55.2<br>(49.5-61.5) | 58.1<br>(50.5-65.8) | <0.001<sup>a</sup> | <0.001<sup>c</sup> |
| Female, n (%) | 132 (60) | 48 (65) | 403 (71) | 72 (60) | 26 (46) | 0.064<sup>b</sup> | <0.001<sup>b</sup> |
| Disease duration median in years (range) | – | 0.3<br>(0.2-0.4) | 9.8<br>(5.7-17.4) | 20.1<br>(14.0-26.7) | 16<br>(8.6-21.2) | – | <0.001<sup>c</sup> |
| EDSS median (range) | – | 1.5 (0-5.5) | 2.0 (0.0-7.5) | 6.0 (1.5-8.0) | 6.0 (2.5-8.0) | – | <0.001<sup>c</sup> |
| Normalized T2 lesion volume (cm<sup>3</sup>) | - | 4.47<br>(± 6.06) | 13.41<br>(± 13.08) | 24.83<br>(± 21.45) | 17.09<br>(± 16.03) | - | <0.01<sup>d*</sup> |
| Normalized brain volume (cm<sup>3</sup>) | 1503<br>(± 101.22) | 1497<br>(± 115.27) | 1470<br>(± 99.51) | 1388<br>(± 83.84) | 1399<br>(± 90.60) | <0.001<sup>d</sup> | <0.05<sup>d**</sup> |

The data represent the absolute numbers and the proportions of the qualitative data, or the median and interquartile range (IQR) for the quantitative data: HCs, healthy controls; MS, multiple sclerosis; CIS, clinically isolated syndrome; RRMS, relapsing-remitting MS; SPMS, secondary-progressive; PP, primary progressive; EDSS, Expanded disability status scale; PwMS, people with MS; a, Wilcoxon signed-rank test; b, Chi-squared test; c, Kruskal–Wallis H test; d, independent two-sample ttest; \*, Significant differences between RRMS vs SPMS and PPMS vs SPMS; \*\*, Significant differences between RRMS vs SPMS, PPMS vs SPMS and RRMS vs PPMS. All statistical analyses were performed after adjusting for age and sex, and applying the ComBat harmonization.

### Network modifications in PwMS

When compared with HCs, the global network graph metrics (Table 2) and local properties were significantly lower in all the PwMS. Moreover, PwMS presented a reduced FA in numerous connections (1686 of 1818 connections, 92.74%), with the strongest differences observed for the intrahemispheric and interhemispheric connections between regions connected to the cingulate, frontal, occipital and dGM structures, such as the right thalamus and right pallidum.

**Table 2.** Global network measures.

|  | **HCS vs PwMS**<br>**p value** | **CIS/RRMS**<br>**vs SPMS**<br>**p value** | **CIS/RRMS**<br>**vs PPMS**<br>**p value** | **SPMS**<br>**vs PPMS**<br>**p value** |
| --- | --- | --- | --- | --- |
| Strength | <0.001 | <0.001 | n.s | <0.001 |
| Betweenness<br>centrality | 0.005 | n.s | n.s | <0.001 |
| Clustering<br>coefficient | <0.001 | <0.001 | n.s | <0.001 |
| Global<br>efficiency | <0.001 | <0.001 | n.s | <0.001 |

HCS, healthy controls; CIS, clinically isolated syndrome; RRMS, relapsing-remitting MS; SPMS, secondary-progressive MS; PPMS, primary progressive MS; n.s., not statistically significant.

### Network changes regarding MS phenotypes

There were similar disruptions to the global and regional properties of CIS (n=73) and RRMS (n=559) networks connections relative to the HCs, affecting 1454 (CIS, 79.98%) and 1484 (RRMS, 81.63%) of the 1818 connections. As no significant differences were found between these phenotypes, we combined them for the analyses (CIS/RRMS). There were significant differences in global (Table 2, *p* values) and local graph network measures between SPMS and CIS/RRMS across all 76 brain network nodes, including strength, clustering coefficient and local efficiency. There was a lower FA in 1452 (79.87%) of the connections associated with the SPMS phenotype, and involving bilateral areas of frontal, parietal, occipital and cingulate cortex, while higher FA values were found for 2 connections from the thalamus and left caudate relative to CIS/RRMS (Figure 1a).

**Figure 1.**
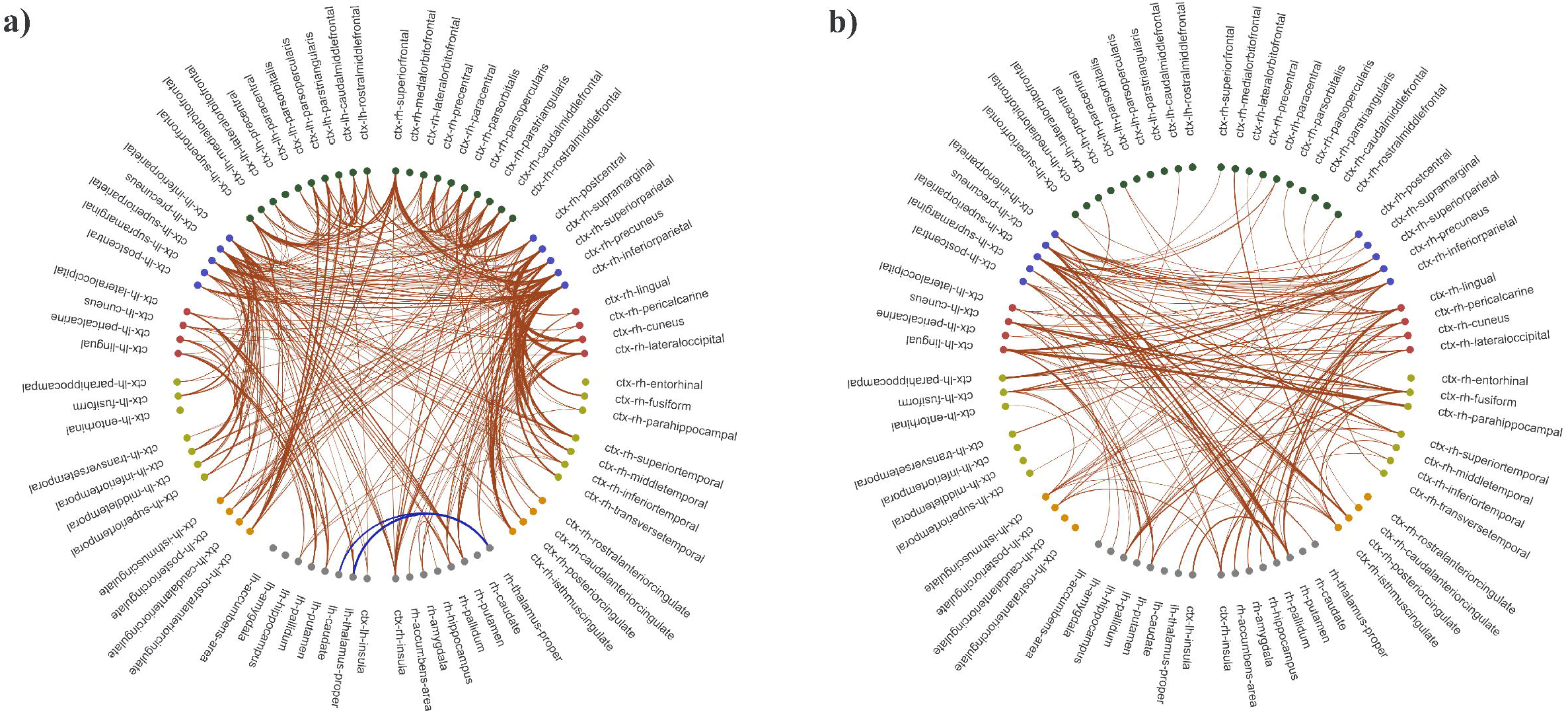
Connectograms comparing the MS phenotypes: a) CIS/RRMS vs SPMS and b) SPMS vs PPMS. Only differences with an effect size above 0.5 are shown (connections with a smaller FA in SPMS than in the other phenotypes are in red and those with a higher FA are in blue): ctx, cortex; lh, left hemisphere; rh, right hemisphere. These connectograms were generated using bokeh from python 3.7 (http://bokeh.org/).

In comparison to CIS/RRMS, there were no statistically significant differences regarding global (Table 2) and local graph measures in PPMS participants. At the same time, the PPMS phenotype was associated with significantly lower FA in 35 connections (1.92%), most of these involving the bilateral frontal and right parietal lobes or the insula, whilst there was also a higher FA at 10 connections within the dGM or the left lateral and medial orbitofrontal cortical regions (Figure 2). When comparing the global network properties of the progressive phenotypes, there were significant differences relative to the SPMS phenotype (Table 2), with a weaker nodal strength at 55.3% of the nodes, lower local efficiency at 60.5% of the nodes and a lower clustering coefficient at 65.8% of the nodes in the SPMS phenotype, whereas there were no significant differences in the betweenness centrality measures. Moreover, there was a lower FA at 303 (16.70%) connections in the SPMS networks relative to the PPMS networks, mainly in the bilateral parietal and occipital cortex or the dGM (Figure 1b).

**Figure 2.**
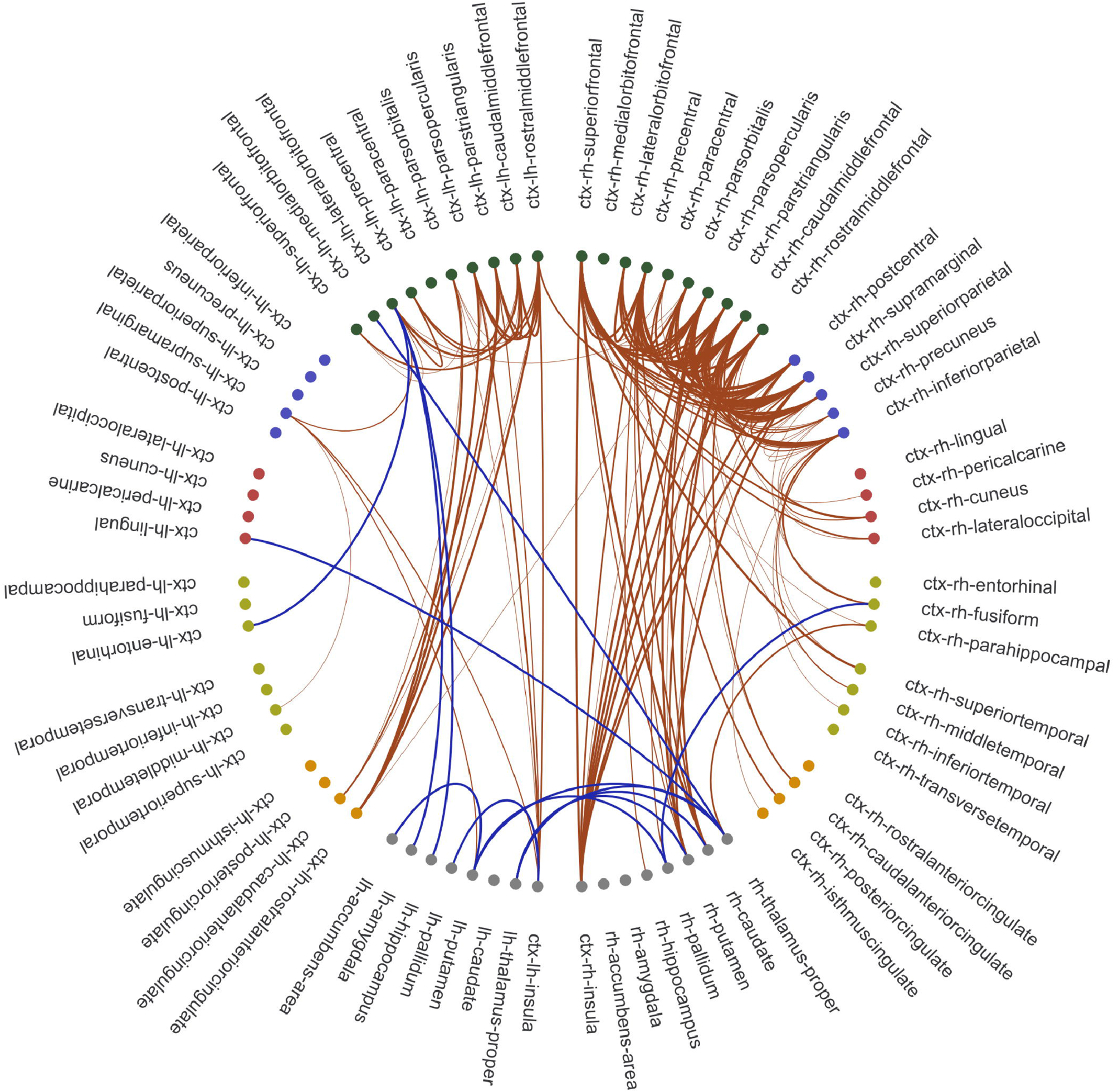
Comparison of the MS connectograms from CIS/RRMS and PPMS. All the significant differences are shown (with a smaller FA in PPMS relative to CIS/RRMS in red and a higher FA in blue): ctx, cortex; lh, left hemisphere; rh, right hemisphere. These connectograms were generated using bokeh from python 3.7 (http://bokeh.org/)

### Classification task according to MS disease and phenotype

The significant differences in the global and regional network characteristics between the phenotypes were used to distinguish PwMS from HCs, and to classify the disease courses. The accuracy of the models to discriminate PwMS from HCs based on FA connectivity matrices (81%) and on local properties (77%) was better than that of the model based on global network graphs (65%: Table 3). The most informative connections to distinguish between PwMS and HCs included dGM structures, such as in the bilateral thalamus, hippocampus, putamen and caudate, or the right pallidum, and cingulate regions.

**Table 3.** Classification measures from super vector machine algorithms to discriminate groups of participants.

|  |  | **HCS vs PwMS** | **CIS/RRMS vs SPMS** | **CIS/RRMS vs PPMS** | **SPMS vs PPMS** |
| --- | --- | --- | --- | --- | --- |
| Global graph metrics | Accuracy | 65 ± 4% | 65 ± 7% | n.a. | 55 ± 8% |
|  | Precision | 60 ± 6% | 60 ± 11% | n.a. | 57 ± 11% |
|  | Recall | 71 ± 2% | 71 ± 3% | n.a. | 54 ± 6% |
| Local graph metrics | Accuracy | 77 ± 3% | 60 ± 5% | n.a. | 56 ± 8% |
|  | Precision | 77 ± 4% | 59 ± 4% | n.a. | 54 ± 8% |
|  | Recall | 77 ± 3% | 62 ± 8% | n.a. | 59 ± 8% |
| FA of connections | Accuracy | 81 ± 4% | 71 ± 7% | 66 ± 4% | 74 ± 5% |
|  | Precision | 81 ± 4% | 66 ± 10% | 63 ± 5% | 66 ± 8% |
|  | Recall | 80 ± 4% | 77 ± 6% | 71 ± 8% | 85 ± 4% |

Classification algorithms could not be applied (n.a.) to metrics that did not show significant differences in the group analyses: HCs, healthy controls; CIS, clinically isolated syndrome; RRMS, relapsing-remitting MS; SPMS, secondary-progressive MS; PPMS, primary progressive MS; ± , standard deviation.

The average accuracy in classifying the MS phenotypes was calculated based on the fraction of significant differences in the connections between CIS/RRMS and SPMS (71%), CIS/RRMS and PPMS (66%), and SPMS and PPMS (74%: Figure 3 and Table 3). As expected, the largest group CIS/RRMS was easier to identify correctly, while the other MS phenotypes were generally more difficult to discriminate with the classification models (see confusion matrix in Figure 3a and 3b). The most important connections in the classification models could be identified (Figure 4), and, the right thalamus, bilateral pallidum and left putamen being particularly important regions to differentiate CIS/RRMS from SPMS. The enhanced FA of the thalamic connections were those with the strongest weights. By contrast, areas of the frontal cortex enabled CIS/RRMS to be differentiated from PPMS, such as the left rostral and right caudal middle frontal cortex. Finally, both the left post-central and parietal cortex (supramarginal and superior parietal) were the regions with the strongest weights to distinguish SPMS from PPMS. Nonetheless, the global and local graph-derived networks performed less accurately than using the FA-weighted connections itself when discriminating the MS subtypes (Table 3).

**Figure 3.**
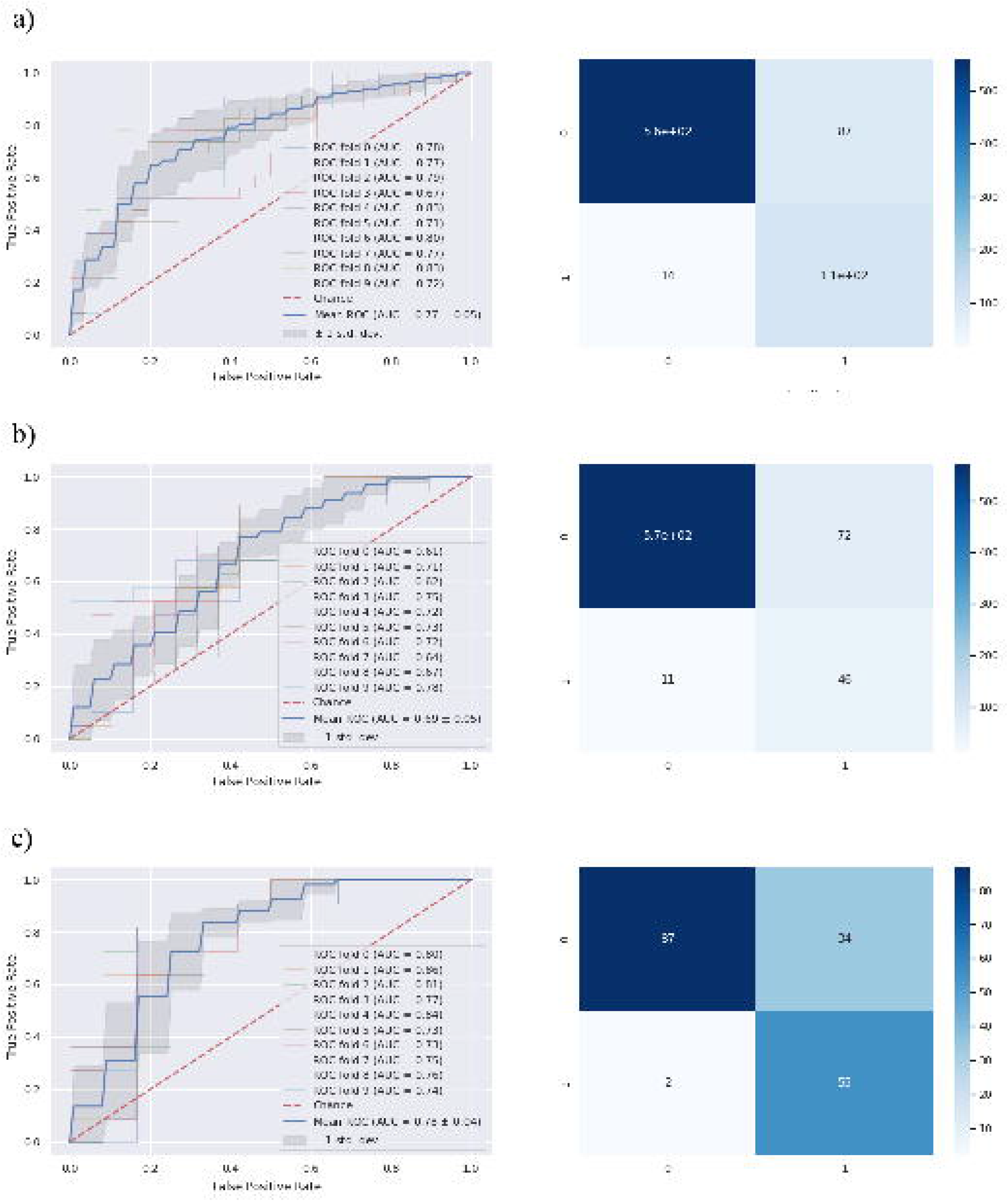
Performance of the classification models for the MS phenotypes and confusion matrices. The receiver operating characteristics (ROC) curves were generated to evaluate the classification performance between a) CIS/RRMS vs SPMS, b) CIS/RRMS vs PPMS and c) SPMS vs PPMS. AUC, Area under the curve.

**Figure 4.**
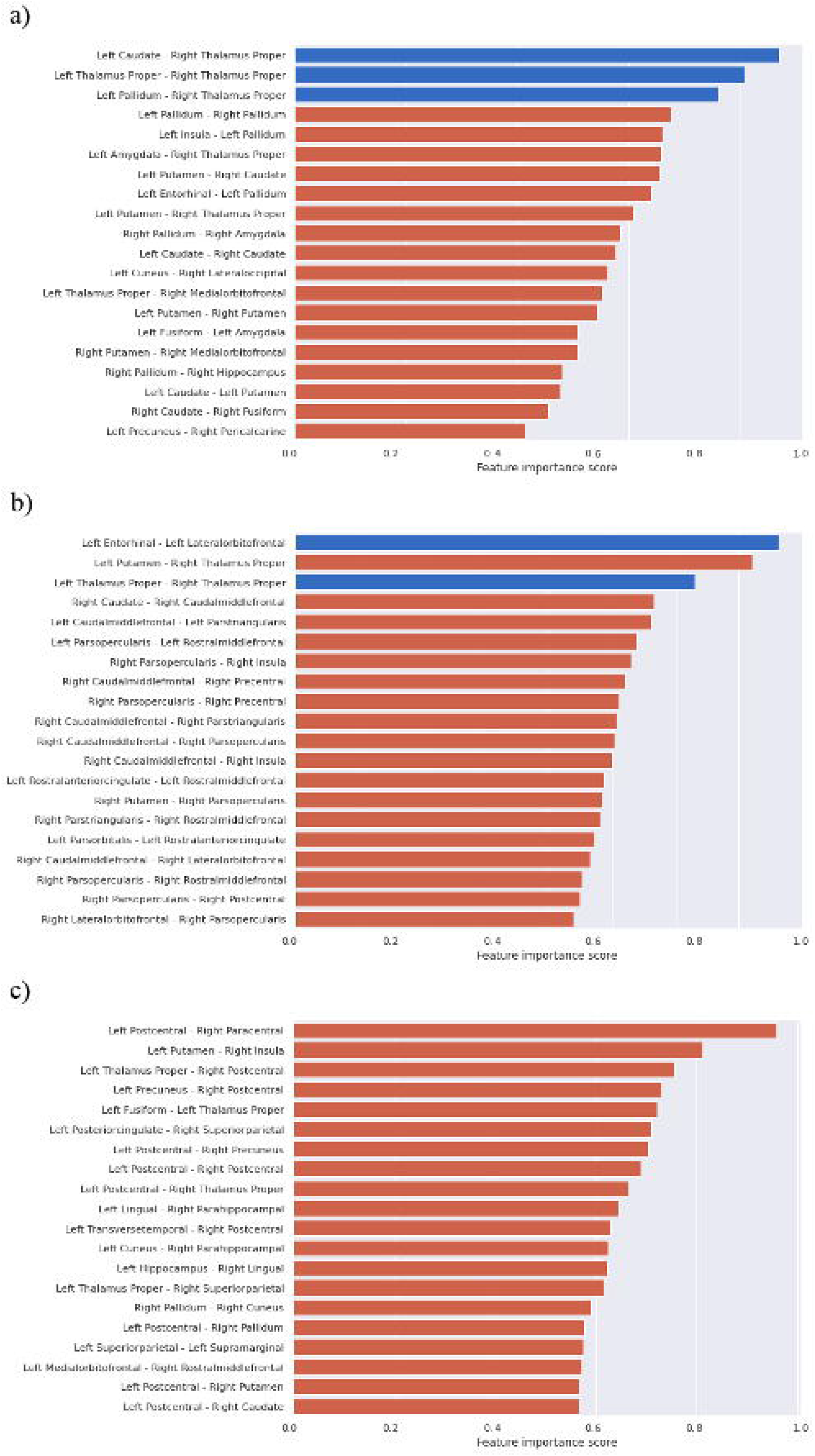
The most important connections established by the ML model to discriminate MS phenotypes based on FA: a) CIS/RRMS vs SPMS; b) CIS/RRMS vs PPMS and c) SPMS vs PPMS. The weights were normalized according to the highest values, with red bars represent lower FA connection weights and blue bars representing higher weights.

## DISCUSSION

In this study, we aimed to assess the clinical relevance of structural network measures in MS. We identified modifications to several components of brain networks that appear to be specific to different MS phenotypes. In PwMS, both the local and global graph properties of the networks and WM connections were altered, which in turn affects global network integrity. The network modifications associated with the CIS/RRMS and PPMS phenotypes are more similar, while SPMS showed stronger abnormalities relative to the other phenotypes. Classification algorithms were able to discriminate the MS phenotypes, with the best accuracy obtained when models use information from FA-weighted connections, especially those regarding thalamic connections, as opposed to those relying on graph metrics.

To address the current lack of knowledge on the value of structural network measures in MS, this multicentre study of advanced MRI data from a large cohort of subjects was designed to assess the diffusion-based structural connectivity characteristics of specific MS phenotypes. In the early stage of the disease, PwMS already express broad modifications to nodal and WM connectivity, triggering global network impairment. In later secondary progressive stages, the network modifications are more extensive, with widespread changes in connectivity associated with the SPMS phenotype ^30^. Indeed, nearly 80% of connections in the SPMS group had a lower FA than in CIS/RRMS, mainly involving bilateral areas of the frontal, parietal and temporal cortex. These results reflect the severe diffuse WM damage at advanced stages of the disease, with higher lesion load, where more normal-appearing white matter damage and more cortical atrophy are usually evident, leading to a more prominent disconnection syndrome ^31,32^. By contrast, the PPMS phenotype differed less from CIS/RRMS, with alterations limited to bilateral frontal and right parietal connections, and with less extensive network changes than in SPMS, as suggested previously ^33^. These findings are concordant with what is commonly observed in PPMS, and with the present results, where the burden of brain lesions is lower and atrophy predominates (Table 1) ^34^. All in all, these results support the idea that these MS phenotypes have a continuum of pathological mechanisms underlying the disease course.

One interesting finding was that the FA of some connections from the thalamus and other dGM structures was higher in SPMS and PPMS than in CIS/RRMS, although the FA values did not reach the values of the HCs. Indeed, atrophy of the thalamus is an early finding in MS ^35^, although silent neurodegeneration at a microstructural level in subcortical projection systems occurs in even earlier stages of the disease ^36^. In progressive forms of MS, changes in neuronal micro-organization occur in conjunction with severe atrophy. Therefore, the significant increase in FA at later phases might be related to tighter packing of WM fibres, which leads to an increase in myelin density and reduce fibre dispersion ^37^.

We analysed the value of features of structural connectivity to differentiate PwMS from healthy individuals and to discern between clinical phenotypes. The resulting ML models demonstrated an accuracy of up to 81% in distinguishing between PwMS and HCs, also identifying the most relevant structural changes in connections from the cingulate cortex and dGM structures like the thalamus and pallidum, areas known for their strong clinical correlations with disability and cognition ^4,38^. The capacity to classify MS phenotypes based on FA-weighted measures was reflected by accuracy values between 66 and 74%, with the greatest accuracy observed in ML models to discern the SPMS phenotype from the others. Nevertheless, the superior discriminative ability of FA-weighted connectivity matrices compared to graph network measures implies that converting the former to the latter may result in the loss of important information. In addition, the most relevant structural connectivity patterns to classify MS subtypes differed depending on the phenotypes compared. Thus, dGM regions were the most relevant to differentiate CIS/RRMS from SPMS, especially these connections involving the thalamus and pallidum. Connections from the parietal cortex (bilateral post-central and superior parietal) and thalamus were the most relevant to differentiate SPMS from PPMS, while tracts involving the frontal cortex (pars opercularis, caudal and rostral middle frontal) were crucial in distinguishing CIS/RRMS from PPMS. Our findings support previous studies on smaller cohorts that reported good discrimination of clinical MS phenotypes using ML-based classification methods and deep learning architectures ^13,14^,^39^, providing evidence of the distinct patterns of WM connectivity in MS.

There are some limitations to this study that should be noted. The dataset was collected at eight MAGNIMS sites that had some heterogeneity in the acquisition protocols, the number of participants and the representation of phenotypes. Thus, we applied the ComBat approach to diminish the individual site effects while preserving the inter-individual FA variability ^16^. The low number of PPMS patients included, which is concordant with the frequency of this phenotype in the disease could have limited the power to detect subtle differences. Quantitative FA metrics may be influenced by the spatial configuration of the tissue, such as the crossing fibres, which can limit its pathological specificity ^40^. Hence, further studies with advanced multicompartmental diffusion methods could help to overcome this limitation and provide a better understanding of the pathological mechanisms underlying the network changes. Finally, future research involving functional connectivity and longitudinal network studies is warranted to gain a more comprehensive understanding of the relationship between structural disconnection and functional reorganization.

In conclusion, structural brain connectivity is disrupted globally in PwMS and differential patterns of regional WM changes are specific to MS phenotypes. As such, CIS/RRMS and PPMS have similar network modifications, while SPMS is associated with more widespread changes in connectivity relative to other phenotypes. Classification methods can distinguish between phenotypes and the largest discriminative value is obtained by considering the integrity of the subcortical connections.

## Supporting information

Supplementary Table 1. Image acquisition parameters at each centre

## Data Availability

All data produced in the present study are available upon reasonable request to the authors

## Acknowledgements

This work was sponsored by the Instituto Carlos III (ISCIII) and co-funded by the European Union through the Plan Estatal de Investigación Científica y Técnica y de Innovación 2015-2024 (PI15/00587 to SL, and AS; PI18/01030 to SL and AS; PI21/01189 to SL and AS), by the Red Española de Esclerosis Múltiple (REEM - RD16/0015/0002, RD16/0015/0003). Part of this work was supported by the German Federal Ministry for Education and Research, BMBF, German Competence Network Multiple Sclerosis (KKNMS), grants 01GI1601I and 01GI0914. The funding bodies had no role in the design and performance of the study; the collection, management, analysis and interpretation of the data; the preparation, revision or approval of the manuscript; and the decision to submit the manuscript for publication. The authors are grateful to the IDIBAPS Magnetic resonance imaging platform for their support. This work was partially developed at the building Centro Esther Koplowitz, Barcelona, CERCA Programme/Generalitat de Catalunya.

## Author Contributions

E.M.-H., E.S and S.LL contributed to the study conception and design. Material preparation, data collection and analysis were performed by E.M.-H. and F.V. The first draft of the manuscript was written by E.M.-H. and S.LL. and all authors commented on further versions of the manuscript. All authors read and approved the final manuscript.

## Data availability statement

Data are available on reasonable request.

