## Supplementary Table 1. Image acquisition parameters at each centre for "Diffusion-based structural connectivity patterns of multiple sclerosis phenotypes"

| *Centre* | *Scanner* | *Sample size (PwMS/HCs)* | *3D-T1 structural* | *Diffusion weighted images* | *EPI correction* |
| --- | --- | --- | --- | --- | --- |
| Hospital Clinic Barcelona | Magnetom Trio 3T SIEMENS | 193 / 45 | 240 sagittal slices with 0.94 mm isotropic voxel size and a 256 × 256 matrix size | 1.5 mm isotropic voxel size, a 154 × 154 matrix size, b value = 1000 s/mm^2^, 60 diffusion encoding directions and a single baseline image acquired at 0 s/mm^2^ | *Field map* |
| Hospital Clinic Barcelona (2) | Magnetom Trio 3T SIEMENS | 116 / 9 | 208 sagittal slices with 0.9 mm isotropic voxel size and a 256 x 256 matrix size | 2 mm isotropic voxel size, a 120 × 120 matrix size, b value = 1500 s/mm^2^, 70 diffusion encoding directions and a single baseline image acquired at 0 s/mm^2^ | *Field map* |
| MS Center, Amsterdam UMC | GE, Signa HDxt | *276 / 69* | 172 sagittal slices with 1 x 0.94 x 0.94 mm voxel size and a 256 x 256 matrix size | 2 x 2 x 2.4 mm voxel size, a 128 × 128 matrix size, b value = 1000 s/mm^2^, 30 diffusion encoding directions and five volumes without directional weighting. | *Synb0* |
| San Raffaele Scientific Institute, Milan | Philips Ingenia CX | *45 / 10* | 204 sagittal slices with 1 mm isotropic voxel size and a 256 x 256 matrix size | 1.9 x 1.9 x 2.3 mm voxel size, a 128 × 128 matrix size, b value = 900 s/mm^2^, 35 diffusion encoding directions and a single baseline image acquired at 0 s/mm^2^ | *T2-weighted image* |
| University Mainz | Magnetom Trio 3T SIEMENS | *45 / 30* | 192 sagittal slices with 1 mm isotropic voxel size and a 256 x 256 matrix size | 2 mm isotropic voxel size, a 128 × 128 matrix size, b-values = 900, 2000 and 3000 s/mm^2^ along 90 diffusion encoding directions, and 7 b = 0 images interleaved for each shell | *T2-weighted image* |
| University of Nottingham, Nottingham | GE 3T MR750 | *31 / 18* | 156 sagittal slices with 1 mm isotropic voxel size and a 256 x 256 matrix size | 2.2 mm isotropic voxel size, a 102 × 102 matrix size, b value = 900 s/mm^2^, 30 diffusion encoding directions and a single baseline image acquired at 0 s/mm^2^ | *T2-weighted image* |
| St. Josef Hospital Ruhr University, Bochum | Philips Achieva 3T | *29 / 29* | 180 sagittal slices with 1 mm isotropic voxel size and a 240 x 240 matrix size | 2.5 mm isotropic voxel size, a 128 × 128 matrix size, b value = 900 s/mm^2^, 32 diffusion encoding directions and a single baseline image acquired at 0 s/mm^2^ | *T2-weighted image* |
| Hospital Vall d’Hebron, Barcelona | Magnetom Trio 3T SIEMENS | 24 / 11 | 128 sagittal slices with 1x1x1.2 mm voxel size and a 256 x 240 matrix size | 2 mm isotropic voxel size, a 122 × 122 matrix size, b value = 1000 s/mm^2^, 30 diffusion encoding directions and a single baseline image acquired at 0 s/mm^2^ | *T2-weighted image* |
| Institute of Neurology, UCL, London | Philips Achieva 3T | 12 / 9 | 180 sagittal slices with 1 mm isotropic voxel size and a 256 x 256 matrix size | 2.3 x 2.3 x 2.5 mm voxel size, a 90 × 90 matrix size, b value = 2000 s/mm^2^, 30 diffusion encoding directions and a four images acquired at 0 s/mm^2^ | *T2-weighted image* |
